# New POCT instrument, GBTsol ICA system for HIV monitoring by CD4 and CD8 enumeration in resource-poor settings

**DOI:** 10.1101/2023.10.12.23296940

**Authors:** Stephe Sung, Hyon-Suk Kim, Jaewoo Song, Jaeyoung Lee, Shabnam Aghamoradi, Seungrim Baek

## Abstract

According to WHO report 39.0 million (33.1–45.7 million) people around the world were living with HIV infection till Dec of 2022. One method that we need for treatment and monitoring of HIV patients is CD4 counting and CD4/CD8 ratio. For this purpose, there are many different laboratory test with technical complexity and in other hand most of them are time consuming, expensive or cannot be used in resource limited places.

This work demonstrates POCT microchip platform for enumerating leukocytes, CD4+ T- lymphocytes, and CD8+ T lymphocytes from whole blood, using fluorochrome-conjugated primary antibodies as a detection method.

This device and its method omits all obstacles for WBC and CD4, CD8 T lymphocytes and it offers fast, cost effective and easy absolute WBC and CD4, CD8 T lymphocytes count for monitoring HIV patients’ immune situation with high accuracy which can be implemented insource limited stings or doctors’ office.

We incubated Phycoerythrin (PE) conjugated primary antibodies specific to CD4 and CD8 antigens to enumerate CD4+ and CD8+ T cells, respectively. Comparison studies were performed with FACS count to evaluate total leukocytes, CD4+T cell number, and CD8+T cell number in whole blood samples for monitoring the immune systems of patients with human immunodeficiency virus (HIV)/AIDS. Statistical analyses for precision, correlation, and agreement were performed. Coefficients of variation (CV) ranging from 0.67% to 12.78%, 0.81 to 13.68%, and 0.29% to 8.33% were obtained for CD4, CD8 and leukocyte recovery respectively. A significant correlation was found between the two assays for CD4 count and CD8 count, with correlation coefficients of 0.90 and 0.91, respectively. Using Bland-Altman plots, a mean bias of 23, 38, and 490 cells/µL (95% CI, n=113) was obtained for CD4, CD8, and total leukocyte count, respectively. These data show that the GBTsol ICA (Immune Cell Analyzer) is comparable to the FACS count platform method for measuring the amounts of CD4 T cells, CD8 T cells, and total leukocytes in blood samples for the purpose of monitoring HIV/AIDS patients with cheap, easy and fast way.

## Introduction

### 1. Importance of CD4 and CD8 enumeration monitoring in HIV/AIDS patients

According to who report 39.0 million (33.1–45.7 million) people around the world were living with HIV infection till Dec of 2022 that most of them live in source limited places.

One method that we need for treatment and monitoring of HIV patients is CD4 counting and CD4/CD8 ratio. For this purpose, there are many different laboratory test with technical complexity and in other hand most of these method is time consuming, expensive or cannot be used in resource limited places. The CD4 cell count has played a central role in the care of HIV-infected children and adults as a measurement of immunosuppression and deciding for antiretroviral therapy (ART). Routine accumulation of CD4 data at diagnosis continues to contribute to treatment priorities and remains critical in identifying late diagnosis [1–3].

One of the reasons behind underutilization of CD4 results is accessibility of clinical centers and centralized clinical laboratories, where currently available technology such as flow cytometry, which is the gold standard method, has been used for 40 years. However, there are many limitations to traditional flow cytometry, including the ability of laser to analyse only one cell at a time, cells must be in suspension to be analysed, highly trained technicians are required, intensive quality control measures are needed, and cells must be viable to be analysed. These requirements have constrained CD4 testing in several resource-constrained settings, most particularly for rural settings where clinical laboratories are not easily accessible [4–8]. Along with the above-mentioned limitations, flow cytometry measurement suffers from a lack of standardization, and reproducible protocols for sample preparation, including RBC lysis, cell staining, gating strategies, and acquisition protocols, have proven difficult to remain constant over multicenter clinical studies [9]. Moreover, patients in many countries lack a unique patient number, making it very difficult to trace test results from different laboratories, especially in patients referred between hospitals and clinics. Long turn-around times for tests sent to central labs delay clinical decision-making and place a significant strain on patients. Conventional tests involve transportation of samples in complex and insufficient transport networks and on lengthy, rough highways. These transport networks are restricted, costly, and often compounded by short sample stability problems. In one study in Mozambique, Malawi, and South Africa, as many as 50% of CD4 test results did not return to clinics for follow up [10]. To address this drawback, health care programs are utilizing POCT for CD4 results to facilitate care for the individual, leading to higher retention in care and reduced loss to follow-up [11]. Point-of-care testing (POCT) are typically used by technicians and other clinical personnel who may not necessarily be adequately trained. Therefore, accuracy, reproducibility, low cost, time effective, sensitivity, and specificity remain the most important factors related to POC technologies. One such POCT instrument is the GBTsol ICA, which has shown high accuracy, reproducibility, low cost, and time effectiveness. This commercial tool of twenty-first century with peculiar CD marker measurements method within multiple maladies provides a solution when and where the absence of similarities in others cannot. Hence, in this study, we focused on the accuracy, reproducibility, and time effectiveness of POC technology (GBTsol ICA) compared to the results of FACS count data. The POC technology (GBTsol ICA) is based on leukocyte separation from whole blood, labelling of all leukocytes with DNA fluorescent stains, and CD marker enumeration (CD4+ and CD8+) using primary antibodies conjugated with fluorescent markers.

### 2. GBTsol ICA system

GBTsol ICA system is composed of three main part. First one is ICA reader machine the second one is cartridge and third part is about reagents and optimized PE conjugated CD marker antibody. Each part is designed in details which has their own consequential rule in experiments’ result.

Inside the ICA reader machine different part works together the hard ware part from cartridge grabber to suction pump, camera, LED and so on. In other hand an associated computer software algorithm is responsible for analysing and converting raw digital image data into counts and percentages which displaying result on device screen in less than 5 minutes and no need for manual cell counting and being worry about human counting errors Fig.1.

**Figure 1.**
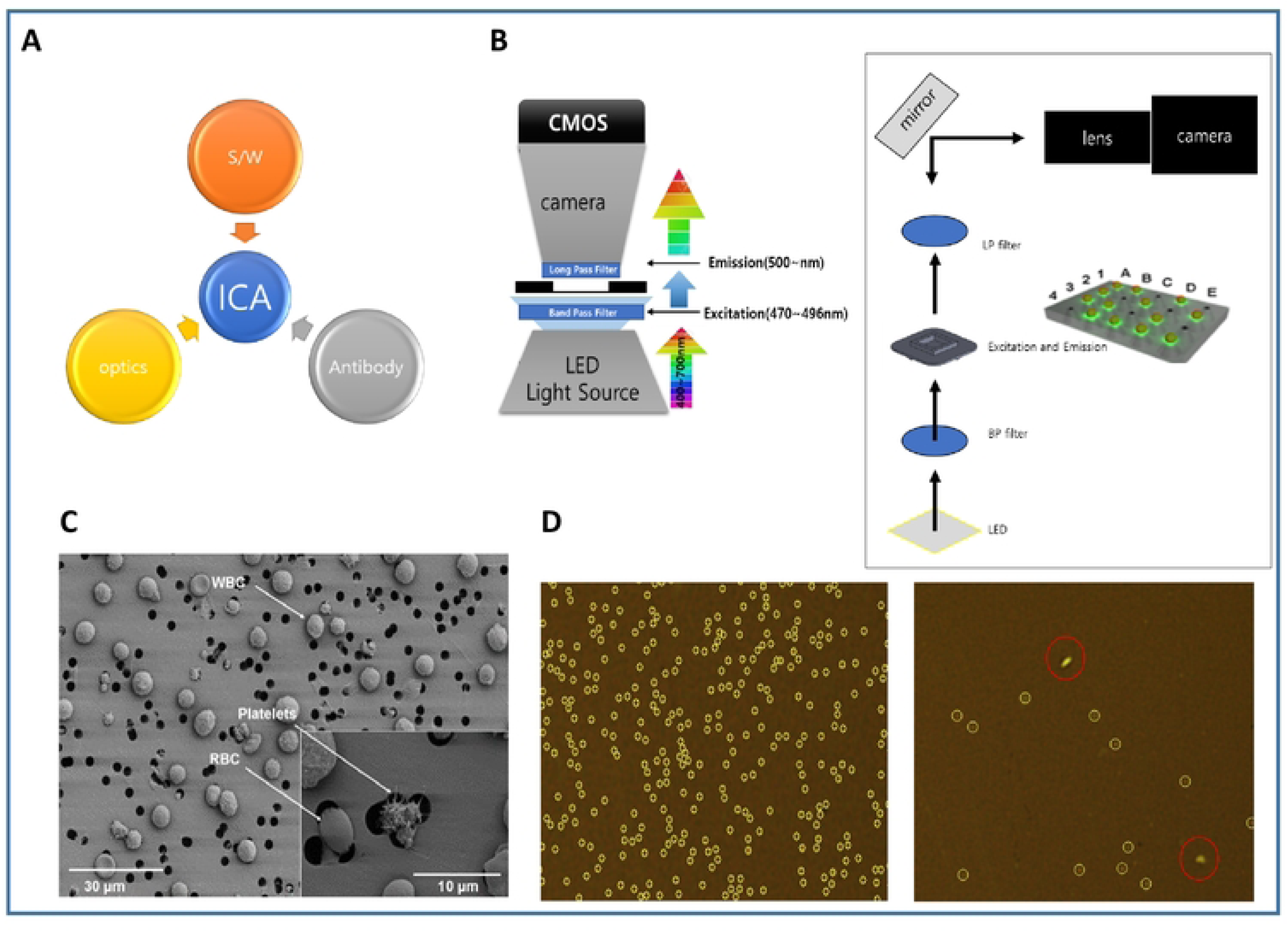
Component of ICA sol system imaging and analyzing. (A) ICA imaging system consist of optic part (Cool LED, Filter,CMOS CAMERA), Software part (graphical user interface, GUI, Image Capture, Analyzing), Biotechnology part (CD and WBC antibody Fluorescence Dye). (B) In optic parts LPF (Long-Pass Filter) filtering out unwanted wavelengths of fluorescence. BPF (Band-Pass Filter) allows signals within a selected range of frequencies to be detect, while preventing signals at unwanted frequencies from getting through. Target cells will bevisible by immunofluorescence antibody markers. (C) Cells more than 3um cannot pass through filter membrane but filter will let the excess dye pass through it during the washing. (D) Software part counts signal and filters the signal with different size or shape from target cell.

The cartridge is disposable and made of three part with specified height and filter membrane with 3um pores, it plays key rule in separating WBC which no need to do RBC lyse or use centrifuge for cell separating which can result in cell lost during experiment.

As CD marker antibody we used mouse IgG Anti-Human CD4-PE from clone RPA-T4 for CD4 marker detection and Mouse IgG Anti-Human CD8-PE from clone OKT-8 for CD8 marker detection. Which are optimized with high purification antibodies and can react with whole blood, no need to lyse RBC and reaction complete just in five minutes. Other reagent and buffers for experiment optimized by designing and processing different protocol to get the best positive signal with low background results wih high accuracy and reproducibility which was validated through testing in Yonsei Medical Center Sinchon Severance Hospital. The data showed close agreement with flow cytometry.

## Methods and materials

Phycoerythrin (PE) conjugated CD4 and CD8 primary mouse antibodies respectively, were centrifuged briefly prior to use. Analytical grade chemicals and reagent grade solvents were used for all buffers and solutions. Dimethyl sulfoxide (DMSO), 0.5% trypsin-ethylenediaminetetraacetic acid (EDTA) solution (10X), and phosphate buffered saline (PBS, pH 7.4) were procured from Sigma Chemical Co. (St. Louis, MO, USA). Fixative and flow cytometry (FC) reagents were acquired from Beckman Coulter (Fullerton, CA). Rainbow microsphere beads were purchased from BD Biosciences, (San Diego, USA). Whole blood samples were obtained by a phlebotomist from anonymous donors at Severance Hospital (Yonsei University, Seoul), in accordance with Institutional Review Board (IRB, 1-2018-009) procedures, by a standard venipuncture technique using EDTA as an anticoagulant in 5mL tubes. Samples were evaluated within 24 hours of collection. Data were obtained from 16/04/2018 to 15/08/2018 .

### 1. Development of biomarker detection kit methodology

Typically, when a patient with HIV/AIDS visits a doctor, blood is collected and sent to the clinical laboratory for CD4 and CD4/CD8 tests with WBC count. Test results may be submitted to the doctor within 24 to 48 hours or up to 1 week depending on number of technicians in laboratory, distance, resources available, and chosen biomarker test. The device presented here is a POC detection kit designed to be used by a health care provider to readily identify and diagnose CD4, CD8 and leukocyte counts.

The GBTsol ICA detection kit consists of the following:

1)A micro-separation filter for entrapment of leukocytes. 2)A tube containing a leukocyte-specific antibody for a specific test. 3)A tube containing reaction buffer for specific reactions (i.e., CD4, CD8, leukocyte counting).4)A tube containing washing solution to remove other proteins non-specifically bound to the micro-separation filter.

Each test using a different kit: GBTsol ICA001 for CD4 count, GBTsol ICA002 for CD8 count, and GBTsol ICA 003 for leukocyte count.

The GBTsol ICA procedure involves five steps. 1) Incubation of sample and respective antibody depending on test i.e., CD4 test, CD8 Test or Leukocyte test. 2)Wetting step- For removal of dust particle or any other unwanted substance from the Cartridge to lower background noise.

3) Sampling step- For dispensing the incubated sample to the GBTsol ICA.

4) Washing Step- For the removal of extra antibody or unconjugated antibody from the cartridge. 5)Processing step- For quantitative result for the CD4, CD8 and Leukocytes respectively.

After the sample is added, the user injects a washing solution, after which the instrument processes the sample automatically. Results can be printed or sent to a storage device.

### 2. Sensitivity test

To establish assay sensitivity, samples of microsphere rainbow beads of different concentrations were dispersed in phosphate buffer saline (pH-7.0) (100, 250, 500, 1000, 1500, 2000, 2500, 3000, 3500, 4000, 4500, 5000 microsphere rainbow beads/µL), and bead counts were performed using GBTsol ICA following the method for cell counting. Counts were repeated three times to determine the average number of microsphere rainbow beads.

### 3. Identification of leukocytes and enumeration of specific blood cell subsets

Human blood samples were collected in EDTA tube and antibodies utilized in these studies were stored at 4∼8oC. Each blood sample (5µL) was mixed with cell-staining solution (5uL) (GBTsol ICA-001), (GBTsol ICA-002) containing fluorescently labeled antibodies targeting CD antigens and incubated at room temperature for 5 min after incubation 990uL of reaction buffer was added. Samples were directly dispensed on the GBTsol ICA cartridge for enumeration of CD markers. GBTsol ICA is a semi-automated and fully quantitative device. After sample injection, the tests were carried out automatically to generate results.

### 4. Flow cytometry analysis

FACSCount flow cytometer (NAVIOUS Ex Flow Cytometer Leukocyte Count) using a protocol provided by the manufacturer was used as a reference method. BD True count tubes (BD Biosciences) were used for determining absolute leukocyte, CD4+, and CD8+ cell counts.

### Data management and analysis

All study data were collected and managed using electronic data capture tools based on our own algorithm and hosted at Glory Biotechnologies Corp., which is a secure, password-protected, web-based application designed to support data capture for research studies. All data were entered once, and each entry was checked for accuracy and was only available to staff directly involved in data entry or analysis. Leukocytes, CD8, and CD4 results were analyzed with Microsoft Excel after being exported directly from the GBTsol ICA device to determine the number of tests performed, invalid test rates, and types of invalid tests. Error was estimated by scatterplot and best-line analyses with linear regression to determine the coefficient of determination (R2), and Bland-Altman analysis was performed to determine systematic bias, limit of agreement (LOA), and imprecision of the GBTsol ICA.

### Image analysis of leukocytes and specific blood cell subsets

Each blood sample was stained with fluorescently labeled antibodies targeting specific CD antigens for 5 min. Fluorescence intensities, cell size and shape were used to identify leukocytes. Specific surface markers were used when classifying leukocyte subpopulation specific fluorescent dye conjugated to antibodies against each specific CD marker.

## Results and Discussion

The need for rapid, reproducible, sensitive, accurate, and non-troublesome tests is of major importance in healthcare. Such a test has obvious advantages over existing tests, as well as other instrument-based systems using microfluidics technology to count cells on cartridges.

### Specimen collection and GBTsol ICA performance

A total of 113 patients from Severance Hospital were tested in four months period. Patient age ranged from 8 to 65 years.

Overall, there were few compromised specimens due to difficulties or errors associated with blood collection. Out of 113 CD4 and CD8 tests performed by GBTsol ICA, 2 tests (1.76%) for CD4 and 2 tests (1.76%) for CD8 failed because of pipetting error or improper sample handling. Out of 113 tests for CD4 and CD8, 1 (0.88%) of each sample were clotted before conducting the CD4 and CD8 tests, respectively (Tables 1 and 2). 0.88% of CD4 tests and 0.88% of CD8 tests were aborted before the test was complete. GBTsol ICA instrument error is likely to occur with improper loading, i.e., volume loaded is either too small or too large (Tables 1 and 2).

**Table 1.**
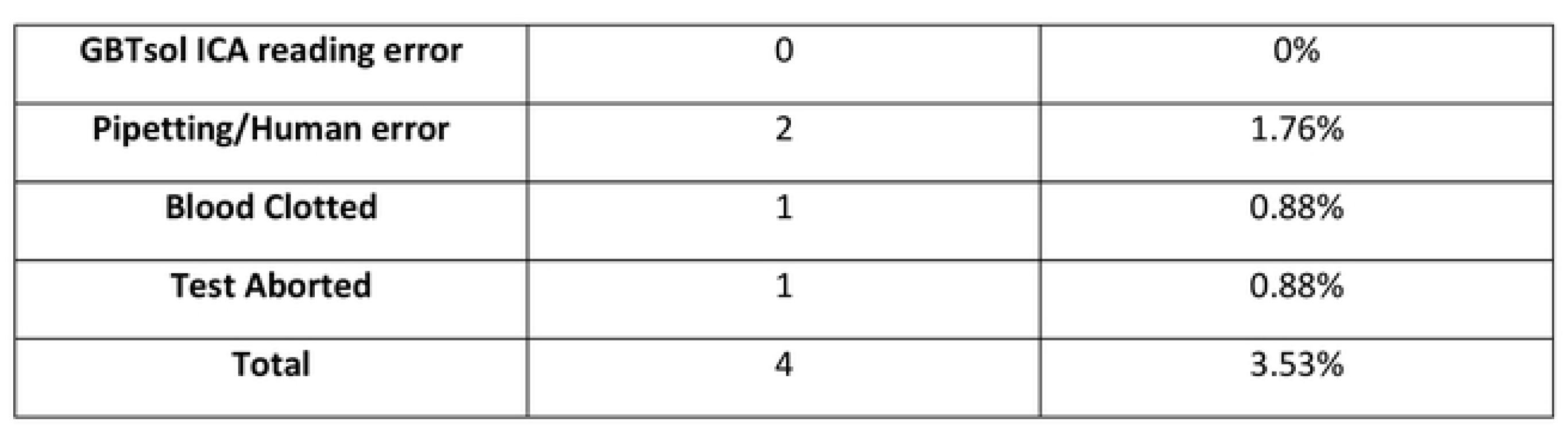
Reason for CD4 results recording failure out of 113 tests performed.

**Table 2.**
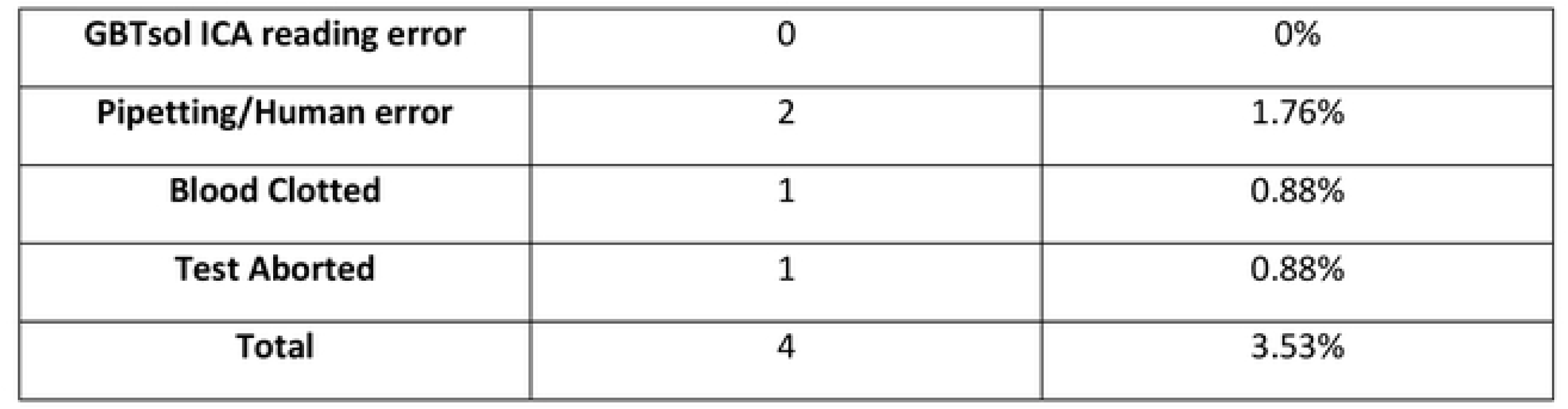
Reason for CDS results recording failure out of 113 tests performed.

However, the GBTsol ICA CD4 and CD8 test cartridges or analyzers could not be totally ruled out as the cause of invalid test results. Operation abortion of GBTsol ICA CD4 and CD8 tests can occur due to mechanical problems or technical problems with the analyser itself or due to testing procedure errors by research personnel. We tried to analyse invalid GBTsol ICA test rates to determine if a particular problem was more responsible for invalid GBTsol ICA tests and concluded that pipetting error or improper sample handling was the major reason for invalid tests. However, out of 113 GBTsol ICA tests for CD4 and CD8, no tests were invalid due to an error message reported by GBTsol ICA respectively. Further investigation suggests that CD4 and CD8 counts less than 100 and greater than 9000, respectively, are not suitable for analysis with GBTsol ICA (data not shown).

However, many existing laboratory tests are time consuming, tedious, and need trained personnel and expensive equipment [12]. POC-CD4 testing is valuable in scale up of ART for HIV care and linkage to treatment in high-risk, resource-restricted settings as conventional flow cytometry measurement of CD4 count generally necessitates samples be sent to a central research facility, which might be off site, with results delayed by 24∼72 hours or even up to 7 days13. POC innovations can lessen such deferrals, allowing faster care. For improvement of testing, high-caliber and low-cost CD4 assays must be accessible for use in resource-restricted settings [12,14]. Such POC instrument and assays must give precise and dependable CD4 results and must be simple to use. Therefore, in this study a novel immunofluorescence test based on biomarker-specific antibodies was used to detect specific CD markers and validated for CD4 and CD8 monitoring for HIV/AIDS patients. This study demonstrates high accuracy and reproducibility in predicting CD4 and CD8 counts using the GBTsol ICA device compared to flow cytometry at Severance Hospital, Yonsei University, Seoul, South Korea.

### Reproducibility and efficiency of GBTsol ICA

To evaluate the detection sensitivity of GBTsol ICA, rainbow beads at concentrations of 100, 250, 500, 1000, 1500, 2000, 2500, 3000, 3500, 4000, 4500, and 5000 rainbow beads/µL were subjected to GBTsol ICA and detected using the device. The calculated detection efficiency was consistently greater than 90% when 100 to 5000 beads were present per 200 µL of rainbow bead sample (Fig. 2). The CV of the different bead concentrations ranged from 2.21 to 5.38%, which adheres to the WHO recommendation < 15% if the count is up to 500 and < 10% if the count is 5000. Accordingly, the GBTsol ICA showed excellent detection efficiency and reproducibility, enabling us to accurately detect CD4, CD8 and leukocyte. (Fig. 4 (A, B), 5 (A, B), 6 (A, B). The GBTsol ICA POC analyzer displayed high reproducibility using normal and low concentrations of beads with coefficients of variation <5%. In this study, the GBTsol ICA POC analyzer overestimated counts compared to the BD FACSCount method in CD4+ T-cell enumeration, in agreement with most studies using capillary or venous blood [15–18]. This overestimation was minimal and is not clinically significant. Differences have been reported in conventional CD4 testing platforms between the BD FACSCount and the BD FACSCalibur, where the mean bias between the two platforms was -76 cells/mm3 (95% CI, LOA - 316.0∼163.0)[19]. Adequate correlation for CD4, CD8, and leukocyte counts between the GBTsol ICA analyzer and the FACSCalibur (0.90, 0.91 and 0.90, respectively) corroborates similar findings [13]. Although a correlation >0.90 was observed between the two platforms for CD4 enumeration, differences were due to variability of instruments settings, antibodies and fluorochromes used, sample volume inputs, and assay procedures and methods.

**Figure 2.**
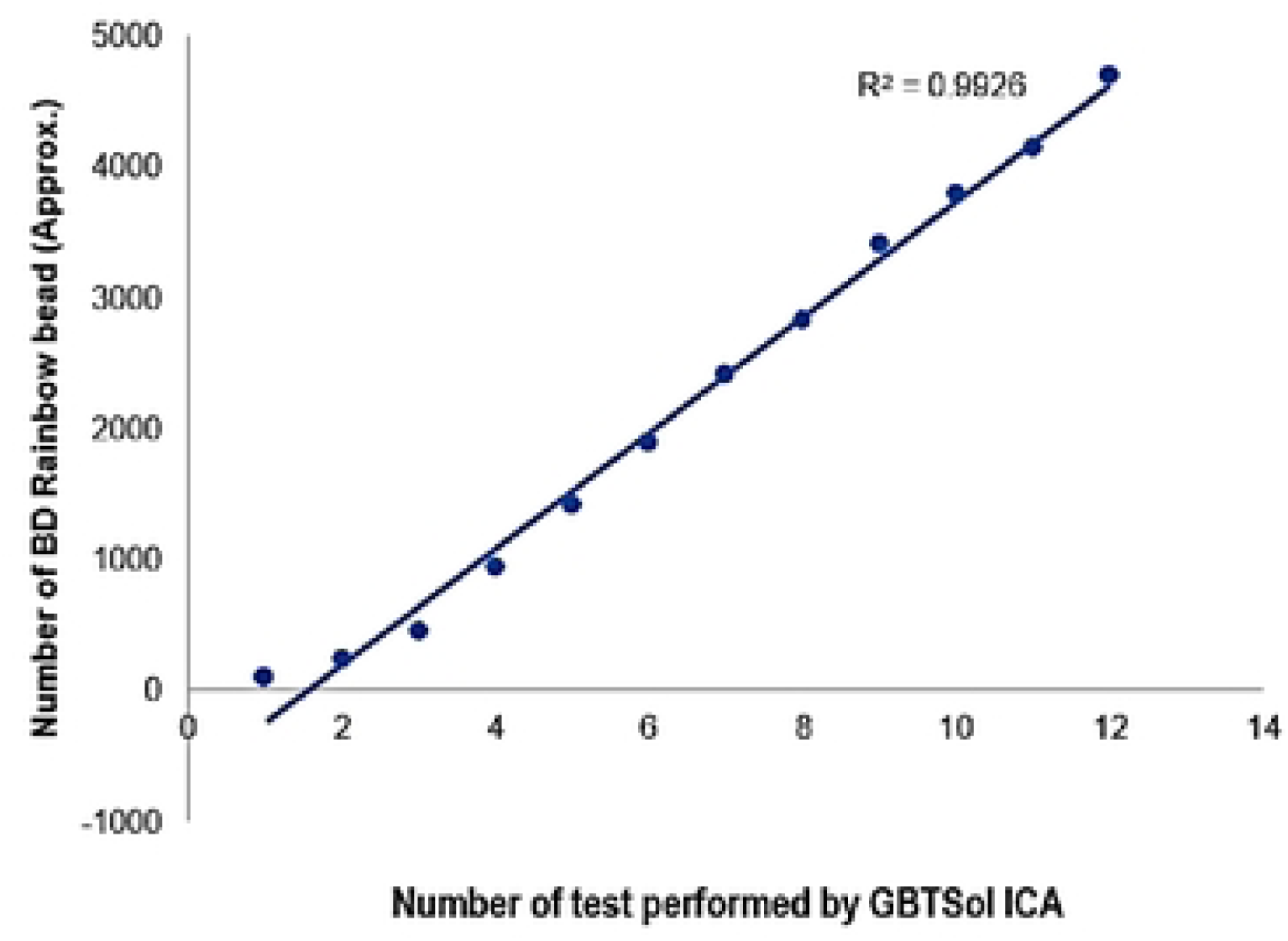
Evaluation of the sensitivity of GBTsol ICA at various target concentrations. A known number of rainbow microsphere beads (100-5000 beads) was added into 1 mL phosphate buffered saline (pH.7.0) and detected using the device integrated with a size-selective micro cavity array. The plot represents the number of cells recovered and the number of tests performed (n=12). CV ranged from 2.28 to 5.38%, and the correlation coefficient (R2) was 0.99.

### WBC count by GBTsol ICA

Leukocyte count was determined by GBTsol ICA for all 113 samples collected within 24 hours and was compared with FACSCount. The recovery of leukocytes was greater than 92% compared to FACSCount, as shown in Fig. 3. The coefficient of determination (R2) was 0.90, the mean bias between the two platforms was 490 cells/µL (95% CI LoA 139.25∼840.84, n=113), and the SD mean was 178.97 (Fig. 4A). These results corroborated to FACSCount results with high accuracy.

**Figure. 3.**
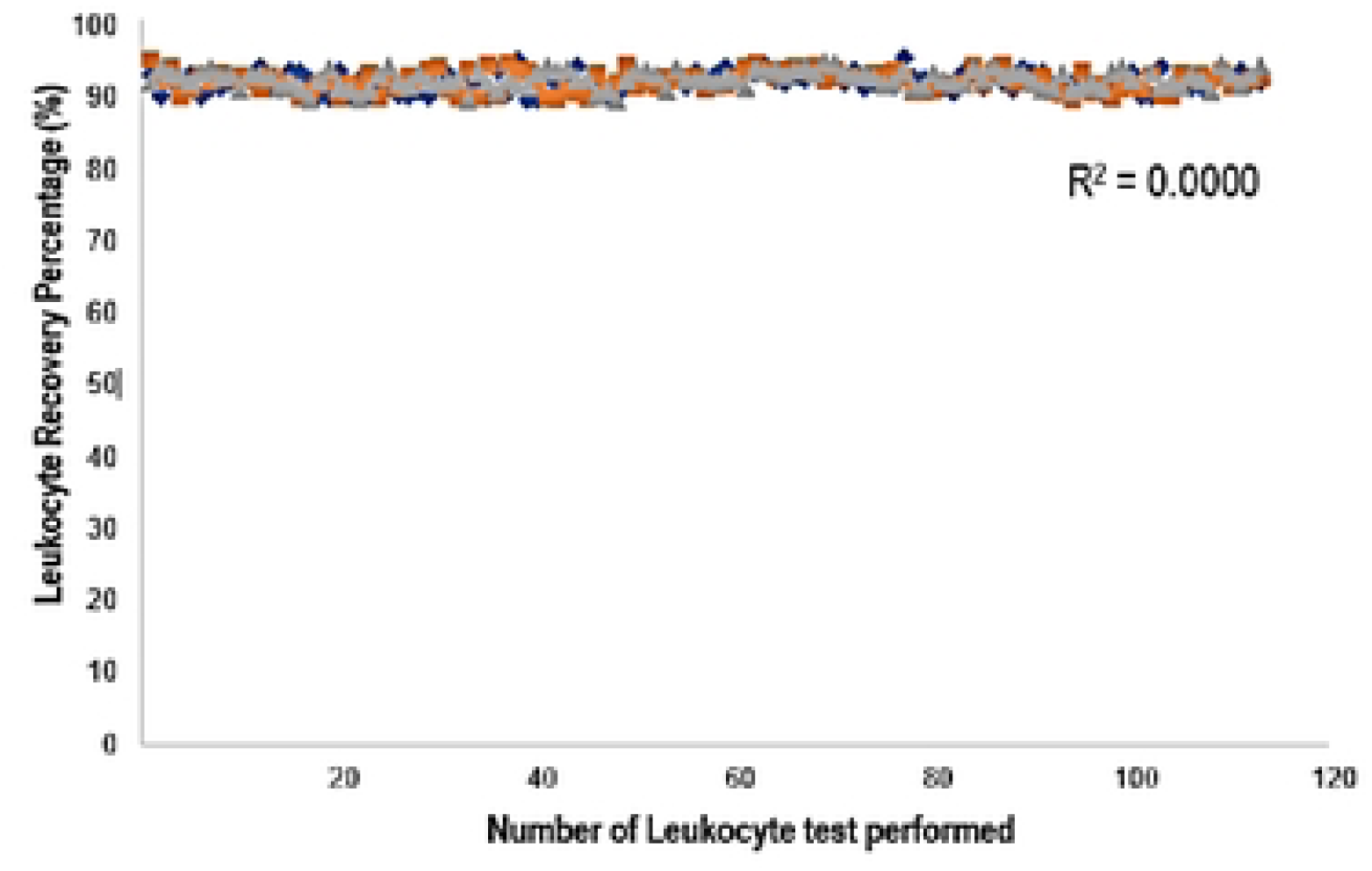
Leukocyte recovery rate from 113 blood samples using the GBTsol ICA device. Blood samples containing 1 uL of whole blood were introduced into theleukocyte-counting device GBTsol ICA. Total leukocytes recovered by the GBTsol ICA were enumerated by an internal computer algorithm using RGB values. Leukocytes were incubated with DNA staining solution from the kit (GBTsol ICA-001). All counts with GBTsol ICA were repeated three times. The average number of leukocytes per microliter of whole blood wascalculated. Thecorrelation value was 0.98,the precision value was <10% (CV%), and the Leukocyte recovery percent was > 92%. Method comparison and correlation studies for total leukocyte counts of 113 human blood samples.

**Figure 4.**
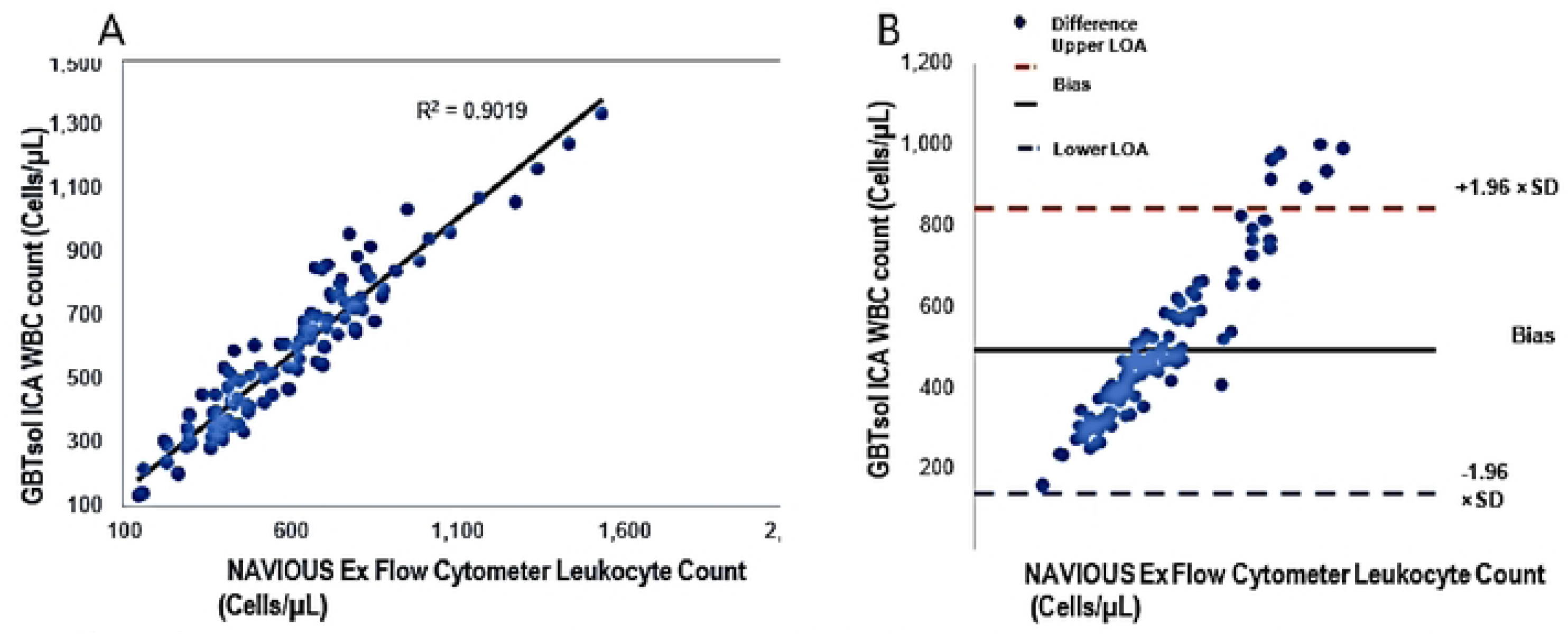
Linear regression and Bland-Altman analysis of GBTsol ICA and FACS count for WBC. In a comparison using venous blood for GBTsol ICA and FACScount, the coefficient of determination (R2) was 0.90 while the mean bias between these two platforms was 490 cells/µL (Cl-95%, LoA 139.25 to 840.84, n=113) andthe SD mean was 178.97. In the Bland Altman plot (Right), the difference between the Oline and the blackline indicatesthebias of GBTsol ICA minus the NAVIOUS ExFlow cytometer.

### CD4 enumeration by GBTsol ICA

A total of 113 patient samples was collected over a period of 4 months under supervision of Yonsei University IRB board members. 200µL of sample containing 1µL of blood was subjected to GBTsol ICA. Next, we tested human whole blood samples with different known CD4+ T-cell counts (as determined by flow cytometry) using the GBTsol ICA. After subtracting the count due to nonspecific binding, we plotted the number of captured cells as a function of CD4 count (Fig. 5A). We obtained a correlation coefficient of 0.90 between our assay and standard flow cytometry. While we intended to use fluorescence only to examine cell capture and noted difficulties in counting individual T-cells in clumps, CD4 count exhibited CVs of 0.67∼12.78%. However, Bland-Altman analysis showed a mean bias of 23 with a 95% CI LoA of -86.47-∼186.16 (n=113) (Fig. 5B).

**Figure 5.**
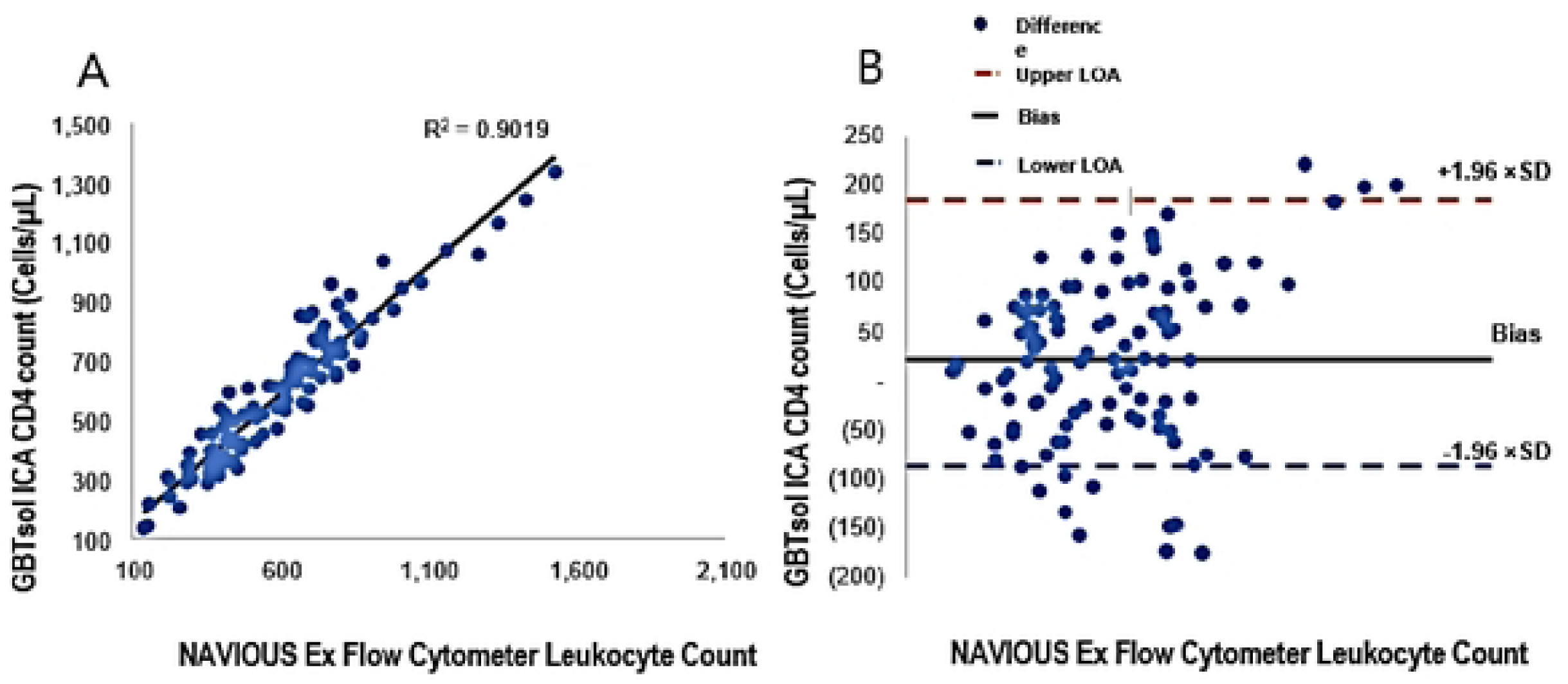
Linear regression and Bland-Altman analysis of GBTsol ICA and FACS count for CD4. In a comparison using venous blood for GBTsol ICA and FACScount, the coefficient of determination (R2) was 0.90 while the mean bias between these two platforms was 23 cells/µL (Cl-95%, LoA -86.47 to 186.16, n=113) and the SD mean was 83.23. In the Bland Altman plot (Right), the difference between the Oline and the black line indicates the bias of GBTsol ICA minus the NAVIOUS Ex Flow Cytometer.

A recent report revealed that same-day POC CD4 testing had no benefit in health outcomes [27]. As supported by other studies [20,21,25,26], we posit the feasibility of use of an existing framework for creation of a small-scale POC research facility to offer tests for staging and pathology of ART.

The overall accuracy of the GBTsol ICA test in samples was observed, and the correlation coefficient was 0.99 to determine the validity of GBTsol ICA. However, in CD4 T-cell count, we observed an overestimation of count. Misclassification, overestimation, or underestimation has been documented in several studies using the PIMA POC analyzer [12,17,20–24].

### Enumeration of CD8 by GBTsol ICA

All samples used for CD4 enumeration were subjected to GBTsol ICA for enumeration of CD8+ T cells. We obtained a correlation coefficient of 0.91 between our assay and standard flow cytometry. In addition, we observed that the number of CD8 count exhibited CVs of 0.81∼13.68% (Fig. 6A). However, the Bland-Altman analysis showed a mean bias of 38 with a 95% CI of LoA of -149.84 ∼ 226.13 (n=113) (Fig. 6B).

**Figure 6.**
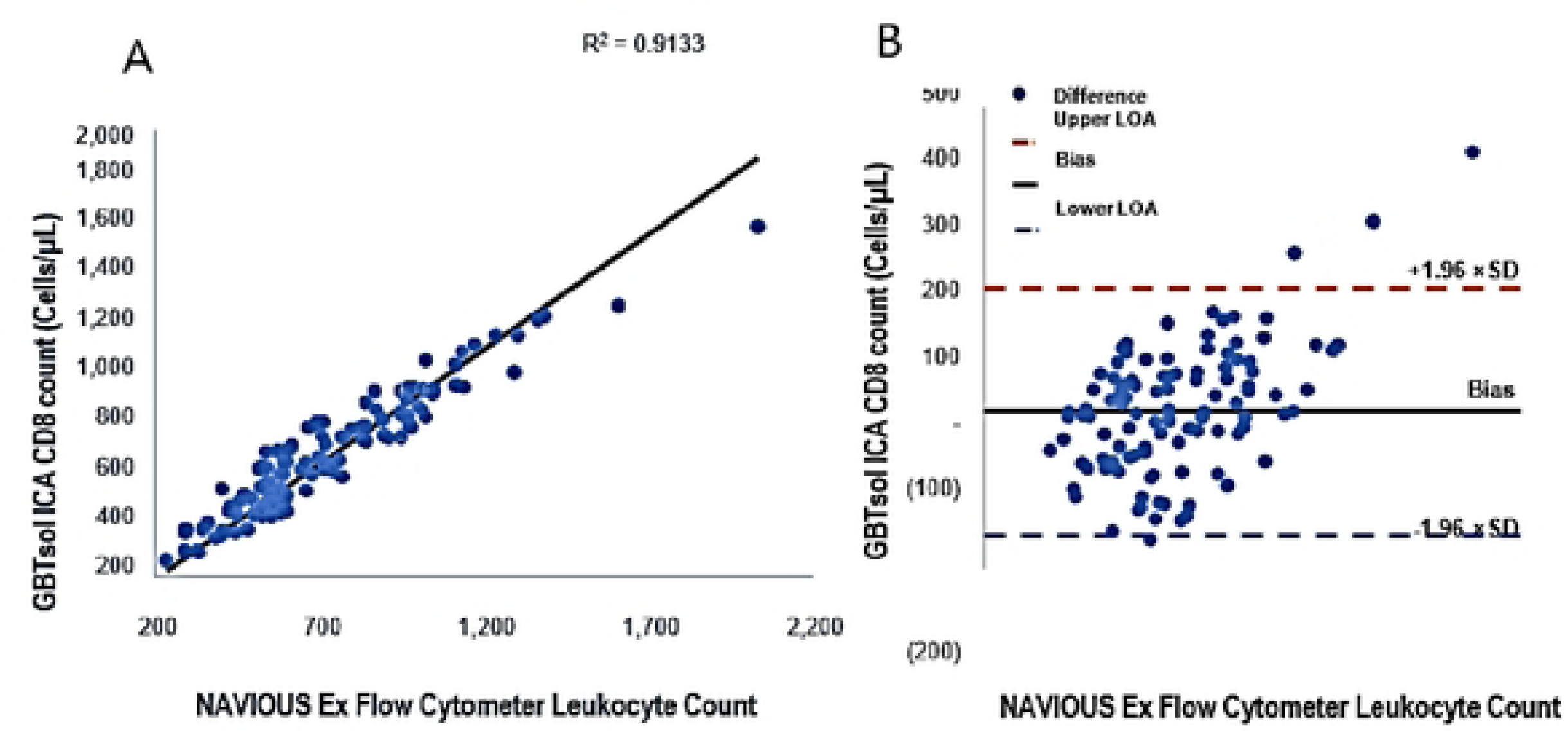
Linear regression and Bland-Altman analysis of GBTsol ICA and FACS count for CDS. In a comparison using venous blood for GBTsol ICA and FACScount, the coefficient of determination (R2) was 0.91 while the mean bias between these two platforms was 38 cells/µL (Cl-95%, LoA-149.84 to 226.13, n=113) and the SD mean was 95.91. In the Bland Altman plot (Right), the difference between the Oline and the black line indicates the bias of GBTsol ICA minus the NAVIOUS Ex Flow Cytometer.

From an operational perspective, use of the GBTsol ICA POC analyzer produced similar results to other studies using venous or capillary blood [12,13,20,22,27]. We experienced test aborted and pipetting/ Human errors of 0.88% % and 1.76% for CD4 and CD8. The operator used in our study was a trained researcher rather than a health professional, such as a nurse or counsellor. Our studies suggest that GBTsol ICA POC is interchangeable with conventional platforms and provides results quite similar to those of PIMA POC technologies [12,16,17,19,20,22,27]. Further testing is needed in communities with HIV/AIDS infection, performed by different health professionals, technicians, and hospitals to verify the coefficient of variation of repeatability and misclassification in favor of under treatment compared to FACSCalibur [19].

Researchers in South Africa observed a median time for patients to return for their CD4 results was 8 days and 7 days in those with ≤ 200 cells/mm3, with a median of 49 days regardless of CD4+ T-cell count from CD4 testing to ART initiation. Use of POC technologies facilitates fast tracking of patients, saves time, and helps in management of HIV treatment, care, and monitoring [28].

## Conclusions

As suggested in a recent systematic review [29], such a resource should streamline administration by limiting patient visits to centers [30], tend to psychosocial issues and boundaries to healthcare [31], enhance counselling and peer support for patients in need [32], optimize the significance of starting and continuing ART if eligible [33], and provide positive wellbeing coaching and encouragement for patients. A family-focused model of coordinated human services consolidating the greater part of the previously mentioned healthcare framework has recently appeared in a comparable populace, yielding high adherence (94%) and maintenance of HIV- 1- positive individuals [34,35]. In this study, we compared a device that enabled highly efficient separation of leukocytes from small amounts of whole blood less than that required by conventional techniques and provided highly accurate enumeration of total numbers of leukocytes and their subsets. We demonstrated that the CD4 and CD8 counts from a few microliters of whole blood using our method had good correlation to flow cytometry analysis.

Thus, our device has potential as an inexpensive yet efficient tool for rapid counting, allowing more detailed leukocyte studies as well as monitoring of HIV/AIDS patients in decentralized sites. The accuracy and simplicity of absolute leukocyte counting offer a range of potential applications in single-cell analysis, including POC diagnostic systems such as human CD marker enumerations for various applications. Previous studies have shown that provision of immediate CD4T+ cell count increased the number of patients receiving care and minimized patient failure to obtain treatment (33%) [11,25,33,36,37].

In conclusion, the overall agreement between FACSCount and the GBTsol ICA analyzer for CD4 T-cell enumeration was acceptable, with a clinically nonsignificant mean bias and high accuracy and reproducibility. We found no significant differences in the results produced from GBTsol ICA compared to those of FACSCount. The GBTsol ICA POC CD4 test exhibited a potential role in CD4 and CD8 T-cell enumeration. This platform can expand access to CD4 testing, particularly in rural settings where needs are currently unmet by existing laboratory testing networks.

## Data Availability

All relevant data are within the manuscript and its Supporting Information files.

## Acknowledgement

This study was approved by the Institutional Review Board (IRB) of Severance Hospital (Seoul, Republic of Korea), an affiliated hospital of Yonsei University Health System (Approval Number: 1-2016-0037). The clinical blood samples were residual blood samples that had been acquired and processed in clinical labs. The acquisition of written consent forms from the patients was waived for using the remaining blood samples after clinical laboratory tests under the IRB. All experiments were conducted in accordance with the relevant guidelines and regulations.

